# Neuropsychiatric adverse events of chloroquine: A real-world pharmacovigilance study using the FAERS database

**DOI:** 10.1101/2020.04.10.20061358

**Authors:** Kenichiro Sato, Tatsuo Mano, Atsushi Iwata, Tatsushi Toda

**Affiliations:** Department of Neurology, Graduate School of Medicine, University of Tokyo; Department of Neurology, Tokyo Metropolitan Geriatric Medical Center Hospital

**Keywords:** COVID-19, chloroquine, adverse events, pharmacovigilance, FAERS, real-world data

## Abstract

**Objectives:** As of the early April 2020, the antimalarial drug, chloroquine, has been approved as an emergency treatment for the coronavirus disease 2019 (COVID-19) in the United States and in Europe. Although infrequent, neuropsychiatric symptoms have been reported in patients who received chloroquine for the treatment of malaria or autoimmune diseases. In this study, we investigated these adverse events (AEs) using a large self-reporting database.

**Materials and Methods:** We conducted a disproportionality analysis for the detection of neuropsychiatric AE signals associated with the use of chloroquine (or hydroxychloroquine), reported to FAERS database between the fourth quarter of 2012 and the fourth quarter of 2019. Adjusted reporting odds ratio (ROR) for the development of each of the neuropsychiatric AEs following the use of chloroquine was calculated using a multilevel model.

**Results:** We included 2,389,474 AE cases, among which 520 cases developed neuropsychiatric AE following the use of chloroquine. Exposure to chloroquine was associated with a statistically significant high reporting of amnesia, delirium, hallucinations, depression, and loss of consciousness, (lower 95% confidence interval of the adjusted ROR > 1), although the degree of increase in their ROR was limited. There was no statistically significant high reporting of any other neuropsychiatric AE, including suicide, psychosis, confusion, and agitation.

**Conclusion:** Current pharmacovigilance study results did not suggest any potential link between the use of chloroquine and an increased risk of suicide, psychosis, confusion, and agitation, which would be informative during the emergency use of chloroquine for the treatment of COVID-19.

## Introduction

The coronavirus disease 2019 (COVID-19) outbreak is a global emergency situation [1], for which the development of drugs and vaccines has been intensively investigated worldwide. In April 2020, the US Food and Drug Administration (FDA) [2] and the European Medicines Agency (EMA) [3] have approved the use of chloroquine and hydroxychloroquine for an emergency treatment of COVID-19, based on their potential effectiveness in this setting [4, 5].

While chloroquine has a long history in the treatment of malaria and autoimmune diseases, its safety and efficacy in the treatment of COVID-19 remain unknown [5]. In the treatment of malaria and autoimmune diseases, chloroquine can cause numerous side effects such as nausea, headache, pruritus, worsening of psoriasis, retinopathy, and cardiac dysfunction. While neuropsychiatric symptoms such as seizure, coma, and psychosis, were also reported in patients who received chloroquine [6, 7], it is still unknown which neuropsychiatric symptoms are directly associated with the use of chloroquine, partly because of their relatively low frequency [8]. Understanding in advance the types of neuropsychiatric symptoms that may occur in patients treated with chloroquine, may be useful, particularly in patients with COVID-19 in intensive care units, where rapid clinical judgement is required.

In the current study, we investigated the link between the use of chloroquine and the occurrence of neuropsychiatric symptoms, using the FDA Adverse Event Reporting System (FAERS) database that is provided by the FDA [9]. Since this database contains a very large number of case reports of drug AEs, it is widely used in identifying potential link between drugs and AEs during post-marketing surveillance of drug safety, [10], despite the risk of bias that is associated with its self-reported data [11].

## Methods

### Data acquisition and preprocessing

This was a retrospective pharmacovigilance study that used the FAERS database. This database contains more than 9 million global case reports of potential adverse events of drugs. On April 1, 2020, we downloaded from the FDA’s website (https://www.fda.gov) patient data that were reported between the fourth quarter of 2012 and the fourth quarter of 2019. The FAERS database contains data tables named ‘DEMO’, ‘DRUG’, ‘REAC’, ‘OUTC’, ‘RPSR’, ‘THER’, and ‘INDI’, of which we mainly used the following 3 tables: 1) ‘DEMO’, which provides the case ID, sex, age, year of event occurrence, country of event occurrence, and the reporter’s type of occupation (e.g., medical doctor, pharmacist, lawyer, consumer); 2) ‘REAC’, which contains all adverse events that are potentially caused by the drug used by each patient, and 3) ‘DRUG’, which includes the name, dose, indication, and date of administration and discontinuation of each drug that is possibly associated with that AE. In the ‘DRUG’ table, the causality assessment of the relationship between each drug and its reported AE is classified by the reporter as ‘primary suspected’, ‘secondary suspected’, ‘concomitant’, or ‘interacting’. In our analysis, we included only reports that were classified as ‘primary suspected’ and ‘secondary suspected’, to reduce the risk of false positive association in deriving our conclusions. In addition, we included only reports that were reported from medical doctor, pharmacist, or other medical staffs, and not from lawyers or consumers. Duplicate reports were excluded such that only the latest version of the report that was obtained from the same case ID was retained.

### Database search

In the FAERS database, the AEs are mainly identified by their Preferred Terms (PTs), as determined by the Medical Dictionary for Regulatory Activities (MedDRA). To search for cases that were associated with chloroquine (including hydroxychloroquine) or mefloquine treatment and presented with any of the AEs of interest, we used the following neurological or psychiatric AE terms, as summarized in Supplemental Table 1: ‘Seizure’ (PT: 10039906), ‘Loss of consciousness’ (PT: 10024855), ‘Confusional state’ (PT: 10010305), ‘Coma’ (PT: 10010071), ‘Stupor’ (PT: 10042264), ‘Somnolence’ (PT: 10041349), ‘Sedation’ (PT: 10039897), ‘Lethargy’ (PT: 10024264), ‘Delirium’ (PT: 10012218), ‘Disorientation’ (PT: 10013395), ‘Hallucination’ (PT: 10019063), ‘Delusion’ (PT: 10012239), ‘Paranoia’ (PT: 10033864), ‘Anxiety’ (PT: 10002855), ‘Anger’ (PT: 10002368), ‘Aggression’ (PT: 10001488), ‘Nervousness’ (PT: 10029216), ‘Irritability’ (PT: 10022998), ‘Mood altered’ (PT: 10027940), ‘Abnormal behavior’ (PT: 10061422), and ‘Restlessness’ (PT: 10038743). In addition, several AE terms were similar, and were therefore treated collectively, based on a single AE category, as follows: ‘Dizziness’ (PT: 10013573) and ‘Dizziness postural’ (10013578) were treated as dizziness, ‘Amnesia’ (PT: 10001949) and ‘Anterograde amnesia’ (PT: 10002711) were treated as amnesia, ‘Altered state of consciousness’ (PT: 10001854) and ‘Mental status changes’ (PT: 10048294) were treated as altered mental status (AMS), ‘Acute psychosis’ (PT: 10001022) and ‘Psychotic disorder’ (PT: 10061290) were treated as psychosis, ‘Agitation’ (PT: 10001497) and ‘Psychomotor hyperactivity’ (PT: 10037211) were treated as agitation, ‘Depressed mood’ (PT: 10012374) and ‘Depression’ (PT: 10012378) were treated as depression, ‘Panic attack’ (PT: 10033664) and ‘Panic reaction’ (PT: 10033670) were treated as panic, ‘Nightmare’ (10029412) and ‘Abnormal dreams’ (10000125) were treated as nightmare, and ‘Suicidal ideation’ (PT: 10042458), ‘Completed suicide’ (PT: 10010144), ‘Suicide attempt’ (PT: 10042464), and ‘Suspected suicide’ (PT: 10082458) were treated suicide.

We included mefloquine as one of our drugs of interest because it is an antimalarial drug that is known to cause neuropsychiatric AEs [12]. We classified each case report based on the following binomial factors: ‘with’ or ‘without’ exposure to the drugs of interest, and ‘with’ or ‘without’ the development of each of the AEs of interest, irrespective of the timing of drug administration or the time of AE development.

### Statistical analyses

All data handling and statistical analyses were performed using R software (version 3.5.1). For each included drug (chloroquine or mefloquine), we calculated the reporting odds ratio (ROR), to assess the potential drug-AE links for chloroquine (or mefloquine) and the reported neuropsychiatric symptoms. The crude ROR was calculated by a 2 × 2 contingency table as described previously [13-15], where all the reports were classified by two factors for each AE term and for each drug, as described above. When the lower limit of the 95% confidential interval (CI) for the adjusted ROR was > 1, the AE was considered to be significantly highly reported following the use of the drug of interest, compared with the report of the same AE following the use of all other drugs.

In calculating ROR, we used a multilevel model, to adjust for covariates in a hierarchical design, as described previously [16, 17]: we incorporated into the model the year of AE occurrence and the country of AE occurrence as random effect terms, to reduce bias in the data. Accordingly, the adjusted ROR was calculated by the following formula [16]:

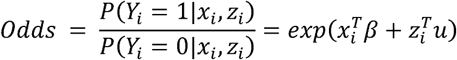

where, *Y*_*i*_ is binomial status for with/without AE of interest in each case *i, β* is the vector of fixed effect parameters, *x*_*i*_ is the covariates matrix for fixed effect, *u* is the vector of random effect parameters, and *z*_*i*_ is the covariates matrix of random effect. Covariates to measure fixed effect include age, sex, binomial exposure to chloroquine (0 if not used, and 1 if used), and binomial exposure to mefloquine (0 if not used, and 1 if used). And covariates to measure random effect (as random intercepts) are the calendar year between 1990 and 2019, and the regional code of the country (e.g., the United States, Great Britain, Canada, Germany, Japan). We used the R package lme4 [18] to perform the above multilevel model analysis.

### Ethics

This study was approved by the University of Tokyo Graduate School of Medicine institutional ethics committee [ID: 11754-(1)]. An informed consent was not required for this type of study. The work was conducted in accordance with the ethical standards laid out in the Declaration of Helsinki, 1964.

## Results

In total, our analysis included 2,389,474 unique case reports. There were 4,336 case reports with exposure to chloroquine, of which 520 (12.0%) reported neuropsychiatric AEs. The number of reported cases with exposure to mefloquine was only 67, of which 28 (41.8%) included neuropsychiatric AEs. Patients treated with chloroquine were predominantly female (3452 females (81.9%) versus 762 males) and the median age of the patients was 56.0 (IQR: 45.0–65.0).

Patients treated with mefloquine were predominantly male (40 males (62.5%) versus 24 females) with a median age of 40.0 (IQR: 27.0–72.0). There were no cases of concomitant exposure to chloroquine and mefloquine.

Figure 1 provides forest plots of the adjusted RORs and the their 95% CI for each neuropsychiatric AE following the use of chloroquine (Figure 1A) or mefloquine (Figure 1B). In the case of chloroquine-associated AEs (Figure 1A), only 5 AE types, including loss of consciousness (LOC) (adjusted ROR, 1.31; 95% CI, 1.03–1.66), amnesia (adjusted ROR, 1.30; 95% CI, 1.02–1.66), delirium (adjusted ROR, 1.58; 95% CI, 1.06–2.34), hallucination (adjusted ROR, 2.12; 95% CI, 1.54–2.92), and depression (adjusted ROR, 1.47; 95% CI, 1.22–1.77) had significantly high reporting (lower limit of the 95% CI > 1). As for mefloquine (Figure 1B), all of the neuropsychiatric AEs, except for somnolence, showed high reporting, with adjusted ROR values higher that 2 in almost all cases.

**Figure 1.**
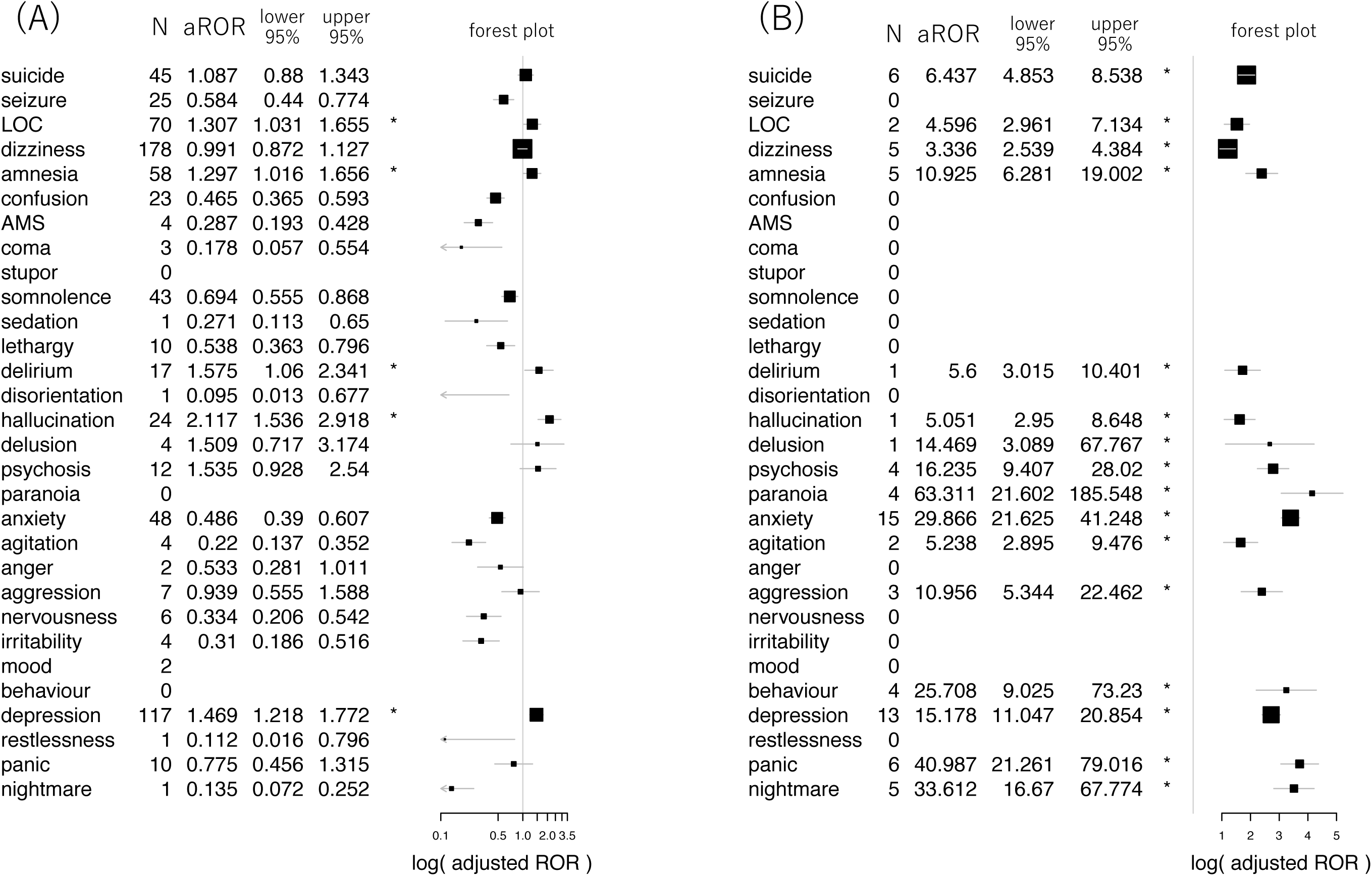
Forest plots, showing the aROR of each AE following the use of chloroquine (A) and mefloquine (B). The number of reports of each AE, the reporting odds ratios that were adjusted based on age and sex for each AE, and their 95% CI are provided in the adjacent tables. Note that the horizontal axis of forest plots is provided in logarithmic scale. For AEs that were not reported after the use of chloroquine or mefloquine (n = 0), the aROR was not calculated and appear as blank. Abbreviations: AE, adverse event; aROR, adjusted reporting odds ratio; LOC, loss of consciousness; AMS, altered mental status, N, number of observations; lower 95%, the lower limit of the 95% confidence interval; upper 95%, the upper limit of the 95% confidence interval.

## Discussion

In this study, we investigated reports of neuropsychiatric AEs that were developed following exposure to chloroquine as a treatment for malaria or autoimmune diseases. The major strength of our study is that it was based on a database that includes global real-world data from a very large number of patients. Our results showed a statistically significant association (albeit marginal) between the use of chloroquine and the reporting of LOC, amnesia, delirium, hallucination, and depression, however, there was no statistically significant association between the use of chloroquine and the reporting of suicide, psychosis, confusion, or agitation, suggesting that there is no link between the use of chloroquine and the increased risk of these AEs. Conversely, mefloquine showed to be associated with statistically significant high reporting of most of the neuropsychiatric AEs, which is consistent with some of the earlier literature reports [12, 19].

Our study has some limitations, resulting from the use of self-reported data [11], and it therefore includes some bias that cannot be eliminated. First, this setting may be associated with a reporting bias, that is, over- or under-reporting of neuropsychiatric AEs following the use of chloroquine. Second, due to the absence of denominators, we were not able to discuss the incidence rate of each neuropsychiatric AE. Third, in our multivariate adjustment, we did not consider the use of concomitant medications or the medical history of the patients, both of which could have potentially contributed to the development or to the worsening of some neuropsychiatric symptoms. Fourth, while suicide was included as one of the AEs in our analysis, it is possible that some of the suicide cases represent suicide by chloroquine overdose, which is known to be cardiotoxic, and not an AE of chloroquine [6]. Fifth, we have not included the timing or the total dose of chloroquine and mefloquine in the analysis. Lastly, it is possible that our final dataset included duplicated cases that should have been excluded.

To conclude, the current pharmacovigilance study, using the FAERS database, did not suggest a potential link between the use of chloroquine and an increased risk of suicide or psychosis. These results provide information that can be essential when the use of chloroquine is considered for the treatment of patients with COVID-19. Since this study was based on a self-reporting database that inevitably contains several biases, cohort studies are needed, to validate these results and to confirm the neuropsychiatric safety of chloroquine.

## Data Availability

The data used in this study is publicly distributed from FAERS database (https://www.fda.gov), which is not owned by the authors.

## Acknowledgments

This study was performed using the FDA Adverse Event Reporting System (FAERS) database that was provided by the FDA. The information, results, or interpretation of the current study do not represent any opinion of the FDA. This study received no funding.

